# Effect of prior traumatic brain injury on Alzheimer’s disease blood biomarkers in Vietnam Veterans

**DOI:** 10.64898/2026.01.13.26344009

**Authors:** Yael Rosen Lang, Agathe Vrillon, Shlomit Pasternak, Ganna Blazhenets, David Soleimani-Meigooni, Gil D. Ravinovici, Michael W. Weiner, Nathan Hantke, Lisa Silbert, Daniel L. Schwartz, Abigail Livny Ezer, Orit Lesman Segev, Ithamar Ganmore, Ramit Ravona-Springer, Kristine Yaffe, Susan M Landau, Magdalena Korecka, Leslie M. Shaw, Renaud La Joie, Raquel C. Gardner

## Abstract

**Importance:** Traumatic brain injury (TBI) is a risk factor for dementia and is known to impact levels of several Alzheimer’s disease (AD) blood biomarkers. Plasma pTau217/ Aβ42 ratio has been reported to be 90% accurate for detection of brain amyloid in civilian cohorts.

**Objective:** To evaluate the accuracy of emerging AD blood biomarkers in Veterans with and without TBI history.

**Design:** We assessed the performance of the FDA-approved plasma pTau217/Aβ42 ratio and plasma levels of pTau217 and Aβ42/40 ratio for detecting brain amyloid-β positivity (e.g., amyloid-PET consensus visual read). We compared biomarkers’ accuracy in Veterans with no TBI (N=93), TBI with loss of consciousness (LOC) 0-5 minutes (N=89), and TBI with LOC >5 minutes (N=90).

**Setting:** Cross-sectional cohort study using existing data and banked plasma from the Alzheimer’s Disease Neuroimaging Initiative Department of Defense (ADNI-DOD) study.

**Participants:** 272 older Vietnam Veterans without dementia (83% cognitively unimpaired, 17% mild cognitive impairment), median (IQR) age 69 (67,72) years, 270/272 male, who had amyloid-PET and concurrently collected banked plasma available for analysis.

**Results:** Amyloid-PET positivity prevalence was 30.5%. Plasma pTau217/Aβ42 ratio was highly accurate (90%) in Veterans with no TBI, but not in Veterans with TBI with LOC 0-5 minutes (78% accuracy, *P*=0.027 vs no TBI) nor in Veterans with TBI with LOC>5 minutes (63% accuracy, *P*<0.001 vs no TBI). Results were similar for plasma pTau217 alone and plasma Aβ42/40 ratio. Results were also similar after excluding Veterans with TBI within the past 10 years, or when amyloid-PET positivity was defined using a quantitative threshold rather than consensus visual read.

**Discussion:** Prior TBI is a modifier of AD biomarkers accuracy in prediction of brain amyloid-PET positivity. Caution is advised in interpreting AD blood test results in this context. Further research is warranted to refine precision AD diagnosis in Veterans and civilians with TBI history.

**Key points:** 

**Question:** What is the accuracy of the plasma pTau217/Aβ42 ratio test for detecting amyloid-PET positivity for Alzheimer’s disease (AD) diagnosis in older Veterans with and without a history of traumatic brain injury (TBI)?

**Findings:** In this cross-sectional study (n=272), plasma pTau217/Aβ42 ratio test accuracy was 90% in Veterans without TBI history, but was significantly lower (63-78% accuracy) in Veterans with TBI history, with lowest accuracy in those with greater TBI severity.

**Meaning:** TBI history and severity is a modifier of AD blood test accuracy in prediction of brain amyloid-PET positivity.AD blood tests should be interpreted with caution in Veterans and civilians with TBI history.

## Introduction

Biomarker-guided diagnosis and treatment of Alzheimer’s Disease (AD) has entered clinical practice. Blood-based biomarkers- loid-beta (Aβ40, Aβ42) and phosphorylated-Tau-217 (pTau217)- are combined into ratios (Aβ42/40 and pTau217/Aβ42) that show promise in detection of cerebral Aβ plaques,^1–3^ the pathognomonic feature of AD targeted by novel amyloid-targeting therapies.^4,5^ While biomarkers offer scalable diagnosis, their accuracy may vary across platforms^6^, subpopulations^7^ and comorbidities.^8^ To optimize diagnostic accuracy and efficiency for individual patients, post-approval validation studies are essential to guide selection and interpretation of AD diagnostic tests, especially in under-represented populations.

In the US there are 16.5 million Veterans, 49% of whom are 65+ years old.^9^ Although Veterans are at high risk for cognitive decline due to high prevalence of dementia risk factors,^10,11^ they are largely underrepresented in AD blood biomarker research. This is problematic because Veterans often have unique factors, such as military-specific exposures, that could affect AD clinical presentation and AD diagnostic test accuracy. Traumatic brain injury (TBI) is common among military Veterans,^12,13^ is a well-established risk factor for dementia in civilians^14^ and Veterans,^15^ and is associated with elevations of AD biomarkers plasma total tau, pTau and Neurofilament light (NfL) in both acute^16–20^ and chronic^21–23^ TBI. Prior TBI is common, occurring in 18-40% of U.S. adults.^12,24,25^ Because TBI is a risk factor for all-cause dementia, individuals presenting for cognitive evaluation may be enriched for prior TBI exposure. However, it is currently unknown if and how TBI history affects the accuracy of emerging AD diagnostic blood tests in Veterans.^26^

In May 2025, the U.S. Food and Drug Administration approved an AD diagnostic blood test, the Fujirebio Lumipulse G pTau217/Aβ42 ratio, in symptomatic adults age 55 years or older with mild cognitive impairment (MCI) or mild dementia. The test is reported to have 90% diagnostic accuracy versus gold-standard tests such as amyloid-β Positron Emission Tomography (Aβ-PET) or cerebrospinal fluid.^2^ We aimed to test AD blood biomarker accuracy versus the gold-standard of Aβ-PET in Vietnam Veterans with and without prior TBI. Our benchmark was 90% accuracy, per expert consensus guidelines for diagnosis and staging of AD with blood biomarkers.^27^ Our goal is to guide appropriate use and interpretation of emerging AD diagnostic blood tests in military Veterans and individuals with prior TBI.

## Methods

### Study design, ethics, and data source

This cross-sectional study leveraged clinical data, Aβ-PET imaging studies, and banked plasma from the completed Alzheimer’s Disease Neuroimaging Initiative Department of Defense (ADNI-DOD) study of older Vietnam Veterans, most of whom had normal cognition.^28^ The study was approved by the Helsinki Committee of Sheba Medical Center, the single IRB of the University of California San Francisco (UCSF), and the US Army Medical Research and Development Command Office of Human Research Oversight. The need for informed consent was waived due to the use of existing de-identified data and samples. Detailed methods of the ADNI-DOD study were previously described. ^28,29^

### Inclusion criteria

We included ADNI-DOD participants who had Florbetapir Aβ-PET imaging suitable for visual and quantitative analysis and concurrently drawn banked plasma.

### Aβ PET acquisition and preprocessing

Florbetapir F18 PET scans were acquired through ADNI-DOD as previously described.^29^ In short, acquisition began 50 minutes post-injection of 10 millicurie and took 20 minutes (4X5 minutes scan). Florbetapir images were re-aligned, averaged, and spatially normalized to MNI-152 space tracer-specific templates.

### Aβ quantification and status

Aβ-PET studies were visually read by one of five trained neurologists from UCSF blinded to demographics, clinical characteristics, and Aβ quantification to determine Aβ status: positive/negative for cortical binding by visual read. Aβ deposition was also quantified usingCentiloids, a standardized measure of Aβ burden.^30^ Centiloids(CL) were derived from a PET-only pipeline, as previously described^31^, as MRIs were not available for all ADNI-DOD participants. A value ≥27 CL was used as a quantitative threshold for amyloid-PET positivity ^31^Scans with discordant visual read and quantitative results were classified by consensus review of all readers.

The final result of this process will be referred to henceforth as Aβ-PET visual read positive versus negative (Aβ-PETv+/-). We also used Standardized uptake value ratio (SUVR) with whole cerebellum reference region, generated as previously described;^29^ SUVR >1.17 was defined as positive.^29^

### TBI exposure definition

Using the Ohio State University TBI Identification Method (OSU TBI-ID),^32,33^ TBI was defined as head injury with loss of consciousness (LOC), post-traumatic amnesia (PTA), or alteration of consciousness (AOC), aligning with previous studies^24,34,35^ and standard clinical criteria.^36,37^

### TBI severity categorization

Veterans with multiple TBIs were categorized according to their most severe TBI. Severity of each TBI was determined based on LOC duration only, aligning with previous studies using the OSU TBI-ID. ^12,38,39^ Because lifetime TBI severity can vary substantially across different cohorts, we examined the distribution of TBI frequency and severity in our cohort and assigned cut-points to divide the population into approximate tertiles of TBI “dose” to support an optimally-powered analysis.

Conversely, the ADNI-DOD study primary publications^28,29^ defined TBI as any head injury with LOC>5 minutes or PTA>5 minutes or AOC>24 hours.

### Cognitive status definition

Cognitive status was assessed using the Telephone Interview for Cognitive Status 11-item questionnaire, an adapted version of the Eight-Item Interview to Differentiate Aging and Dementia, and the Clinical Dementia Rating (CDR) interview. Final diagnosis of MCI versus normal cognition was previously made by an ADNI-DOD study clinician^29^ and may differ from CDR global categorization.

### Plasma biomarkers

Aβ40, Aβ42 and pTau217 were measured with the Fujirebio assays on Lumipulse G-1200,and with the Quanterix Simoa Alzpath pTau-217 CARe Advantage kit and Quanterix Simoa 3-plex Advantage kit on Simoa HD-x platform. Glial fibrillary acidic protein (GFAP) is a marker of neurodegeneration and central nervous system trauma.^40,41^ Neurofilament light (NfL) is a biomarker for axonal damage, due to neurodegeneration,^42^ inflammation, trauma or cerebrovascular disease.^43^ Both GFAP and NfL are associated with AD severity.^44,45^ They were measured using the Quanterix Simoa 2-plex kit on the HD-x platform. Before analysis, the samples were stored at -80°C in 0.5 mL aliquots. Two separate aliquots were used to analyze all the biomarkers. Importantly, per standard ADNI biospecimen processing procedures,^46^ all plasma for a participant time-point was first pooled into a single collection tube prior to being aliquoted, thus mitigating potential variation in biomarker levels across individual aliquots.

### Statistical analysis

#### Power considerations

The reported accuracy of plasma pTau217/Aβ42 ratio for Aβ PET positivity detection is 90%.^1,2^ For similar accuracy, with α=0.05 we are powered to estimate the accuracy for Aβ positivity detection in each TBI subgroup (n≈90) with a confidence interval width of 12.4%, or 19% for 70% accuracy.

#### Analysis

We described our cohort’s demographic and clinical characteristics, stratified by TBI severity tertiles, with median and interquartile range (IQR), frequencies and percentages, and compared them using Kruskal-Wallis rank-sum tests, Fisher’s exact tests and chi-square tests. We tested pTau217, Aβ40/42 and pTau217/Aβ42 analyzed by Fujirebio Lumipulse and Quanterix Simoa assays for Aβ positivity detection, compared to a gold-standard of an Aβ-PET visual read, using logistic regression models and Receiver Operating Characteristic (ROC) curves. Biomarker/biomarker ratios were log-transformed to satisfy model assumptions. For each biomarker/biomarker ratio we chose a threshold that optimized overall accuracy,^27^ and then derived accuracy, Area Under the ROC Curve (AUC), negative predictive value (NPV), positive predictive value (PPV), sensitivity and specificity for subgroups of TBI using the ROC curve and confusion matrices.

Biomarker ratio pTau217/Aβ42 has FDA-approved dual diagnostic cut-offs, which classifies results as AD negative (≤ 0.00370), positive (≥ 0.00738) or indeterminate (in between). The same methods were applied to estimate test performance in those classified as positive or negative according to dual cut-offs. We compared pTau217/Aβ42 ratio accuracy between TBI groups (with both single and dual cutoffs) using pairwise 2-tailed 2-proportions z-tests. We plotted the distribution of biomarkers NfL and GFAP by Aβ positivity and TBI group, and tested the difference between Aβ-PETv+ and Aβ-PETv- in each TBI group using Wilcoxon-Mann-Whitney tests.

#### Sensitivity analyses

Since a recent TBI may affect AD biomarkers, we repeated these analyses excluding participants who reported a TBI in the past 10 years. We also tested performance by cognitive status (unimpaired vs MCI) and also using quantitative SUVR in lieu of consensus visual read.

Statistical analyses were performed in R 4.4.2.^47^ The “pROC” package was used for ROC analysis. A two-sided *P*-value <0.05 was considered statistically significant.

## Results

Worst TBI severity and TBI frequency are described (**Supplementary Table S1**). To achieve TBI tertiles we implemented cut-points at no TBI, TBI with LOC 0-5 minutes, and TBI with LOC>5 minutes. Prevalence of >2 TBIs was 8% in the TBI with LOC 0-5 minutes group and 19% in the TBI with LOC>5 minutes group.

In our cohort (n=272), median age (range) was 68.9 years (61-85) and only two were women, both cognitively unimpaired (**Table 1**). Amyloid-PET positivity prevalence was 30.5% overall, 18% in no TBI, 27% in TBI with LOC 0-5min and 47% in TBI with LOC>5min (*P*<0.001). Increasing TBI severity was associated with higher prevalence of MCI (*P*=0.016), and Aβ-PET positivity regardless of how it was defined: visual read (*P*<0.001), SUVR (≥1.17; *P*=0.027), or centiloids (≥27; *P*=0.002). Age, race, education and APOE ε4 status were similar across groups.

**Table 1.** Demographics and baseline characteristics.

| Median (IQR), n (%) | Overall<br>N = 272<sup>1</sup> | None<br>N = 93<sup>1</sup> | TBI with<br>LOC 0-5min<br>N = 89<sup>1</sup> | TBI with<br>LOC >5min<br>N = 90<sup>1</sup> | p-value<sup>2</sup> |
| --- | --- | --- | --- | --- | --- |
| **Age** | 68.9 (66.7, 72.1) | 68.2 (65.9, 71.3) | 69.0 (67.3, 72.2) | 69.3 (67.3, 72.7) | 0.091 |
| **Male gender** | 270 (99%) | 93 (100%) | 87 (98%) | 90 (100%) | 0.11 |
| **Race** |  |  |  |  | 0.7 |
| White | 231 (85%) | 77 (83%) | 79 (89%) | 75 (83%) |  |
| Black | 20 (7.4%) | 8 (8.6%) | 6 (6.7%) | 6 (6.7%) |  |
| Other | 21 (7.7%) | 8 (8.6%) | 4 (4.5%) | 9 (10%) |  |
| **BMI** | 29.7 (26.6, 32.9) | 30.1 (27.8, 33.1) | 29.0 (25.3, 32.0) | 29.9 (26.9, 32.7) | 0.2 |
| **Years of education** | 15 (13, 17) | 15 (13, 18) | 16 (14, 16) | 14 (13, 17) | 0.6 |
| **APOE ε4 carrier** | 68 (26%) | 22 (24%) | 26 (31%) | 20 (24%) | 0.5 |
| (Missing) | 13 | 1 | 5 | 7 |  |
| **Cognitive status** |  |  |  |  | **0.016** |
| Normal | 227 (83%) | 85 (91%) | 74 (83%) | 68 (76%) |  |
| MCI | 45 (17%) | 8 (8.6%) | 15 (17%) | 22 (24%) |  |
| **Baseline Clinical Dementia Rating** |  |  |  |  | 0.6 |
| 0 | 181 (70%) | 66 (73%) | 57 (70%) | 58 (66%) |  |
| 0.5 | 79 (30%) | 24 (27%) | 25 (30%) | 30 (34%) |  |
| (Missing) | 12 | 3 | 7 | 2 |  |
| **Baseline ECog-39** | 59 (48, 77) | 53 (47, 67) | 62 (52, 77) | 63 (48, 84) | **0.043** |
| **Post-traumatic stress syndrome** |  |  |  |  | **<0.001** |
| Negative | 110 (42%) | 49 (54%) | 30 (35%) | 31 (36%) |  |
| Indeterminate | 117 (45%) | 41 (46%) | 47 (55%) | 29 (34%) |  |
| Positive | 35 (13%) | 0 (0%) | 9 (10%) | 26 (30%) |  |
| (Missing) | 10 | 3 | 3 | 4 |  |
| **Cardiovascular disease** | 220 (81%) | 73 (78%) | 73 (82%) | 74 (82%) | 0.8 |
| **TBI in the last 10 years** | 26 (15%) | 0 (NA%) | 14 (16%) | 12 (13%) | 0.7 |
| (Missing) | 93 | 93 | 0 | 0 |  |
| **TBI timing** |  |  |  |  | **<0.001** |
| TBI in the past year | 15 (5.5%) | 0 (0%) | 9 (10%) | 6 (6.7%) |  |
| TBI 1-10 years ago | 11 (4.0%) | 0 (0%) | 5 (5.6%) | 6 (6.7%) |  |
| TBI over 10 years ago | 153 (56%) | 0 (0%) | 75 (84%) | 78 (87%) |  |
| No TBI | 93 (34%) | 93 (100%) | 0 (0%) | 0 (0%) |  |
| **Years since TBI (where applicable)** |  |  |  |  | 0.8 |
| Median (Q1,Q3) | 46 (34,49) | NA (NA,NA) | 47 (38,49) | 46 (26,49) |  |
| (Min - Max) | (0 - 66) | (Inf - -Inf) | (0 - 66) | (0 - 65) |  |
| (Missing) | 93 | 93 | 0 | 0 |  |
| **Aβ PET visual read** |  |  |  |  | **<0.001** |
| Amyloid+ | 83 (31%) | 17 (18%) | 24 (27%) | 42 (47%) |  |
| Amyloid- | 189 (69%) | 76 (82%) | 65 (73%) | 48 (53%) |  |
| **Centiloids** | 12 (1, 27) | 10 (1, 21) | 7 (-3, 22) | 16 (3, 36) | **0.021** |
| **Aβ+ by centiloids (>27)** | 67 (25%) | 16 (17%) | 17 (19%) | 34 (38%) | **0.002** |
| **Aβ+ by centiloids' dual threshold** |  |  |  |  | 0.085 |
| < 10 (negative) | 124 (46%) | 45 (48%) | 48 (54%) | 31 (34%) |  |
| 10 - 40 (indeterminate) | 104 (38%) | 35 (38%) | 30 (34%) | 39 (43%) |  |
| > 40 (positive) | 44 (16%) | 13 (14%) | 11 (12%) | 20 (22%) |  |
| **PET Standardized Uptake Value Ratio (SUVR)<sup>3</sup>** | 1.09 (1.04, 1.16) | 1.09 (1.04, 1.14) | 1.07 (1.02, 1.15) | 1.12 (1.05, 1.20) | **0.025** |
| (Missing) | 3 | 1 | 1 | 1 |  |
| **Aβ+ by SUVR (> 1.17)<sup>2</sup>** | 64 (24%) | 17 (18%) | 17 (19%) | 30 (34%) | **0.027** |
| (Missing) | 3 | 1 | 1 | 1 |  |
| <sup>1</sup> Median (Q1, Q3); n (%) |  |  |  |  |  |
| <sup>2</sup> Kruskal-Wallis rank sum test; Fisher's exact test; Pearson's Chi-squared test |  |  |  |  |  |
| <sup>3</sup> SUVR derived using methods described by Wiener et al., 2023 |  |  |  |  |  |
| Abbreviations: Aβ= amyloid-β, Aβ PET= Aβ positron emission tomography, APOE4=Apolipoprotein E gene, ECOG-39= Everyday cognition-39 item questionnaire, LOC= Loss of consciousness, SUVR= Standardized uptake value ratio, TBI=Traumatic brain injury. |  |  |  |  |  |

Lumipulse pTau217/Aβ42 ratio had an accuracy of 90% (CI: 84%-96%) in participants without TBI history (**Table 2**). However, all 3 biomarkers showed lower accuracy, PPV and sensitivity in participants with TBI exposure, with a dose-response trend for worse accuracy with increasing TBI severity (**Figure 1**). Differences in accuracy were statistically significant for pTau217/Aβ42 ratio: 90% in no TBI vs 78% in TBI with LOC 0-5min (*P*=0.027) and no TBI vs 63% in TBI with LOC>5min, (*P*<0.001) and for pTau217 (87% vs. 79% and vs. 62%, *P*=0.14 and *P*<0.001).. Sensitivity and PPV were lower in TBI groups while NPV and specificity were overall similar across TBI groups (**Table 2**). Biomarkers analyzed by the Quanterix Simoa platform showed very similar results (**Table 2**), with maximal accuracy of 89% for pTau217 and 88% for pTau217/Aβ42 in the no TBI group and much lower accuracy (≤80%) in both TBI groups.

**Figure 1:**
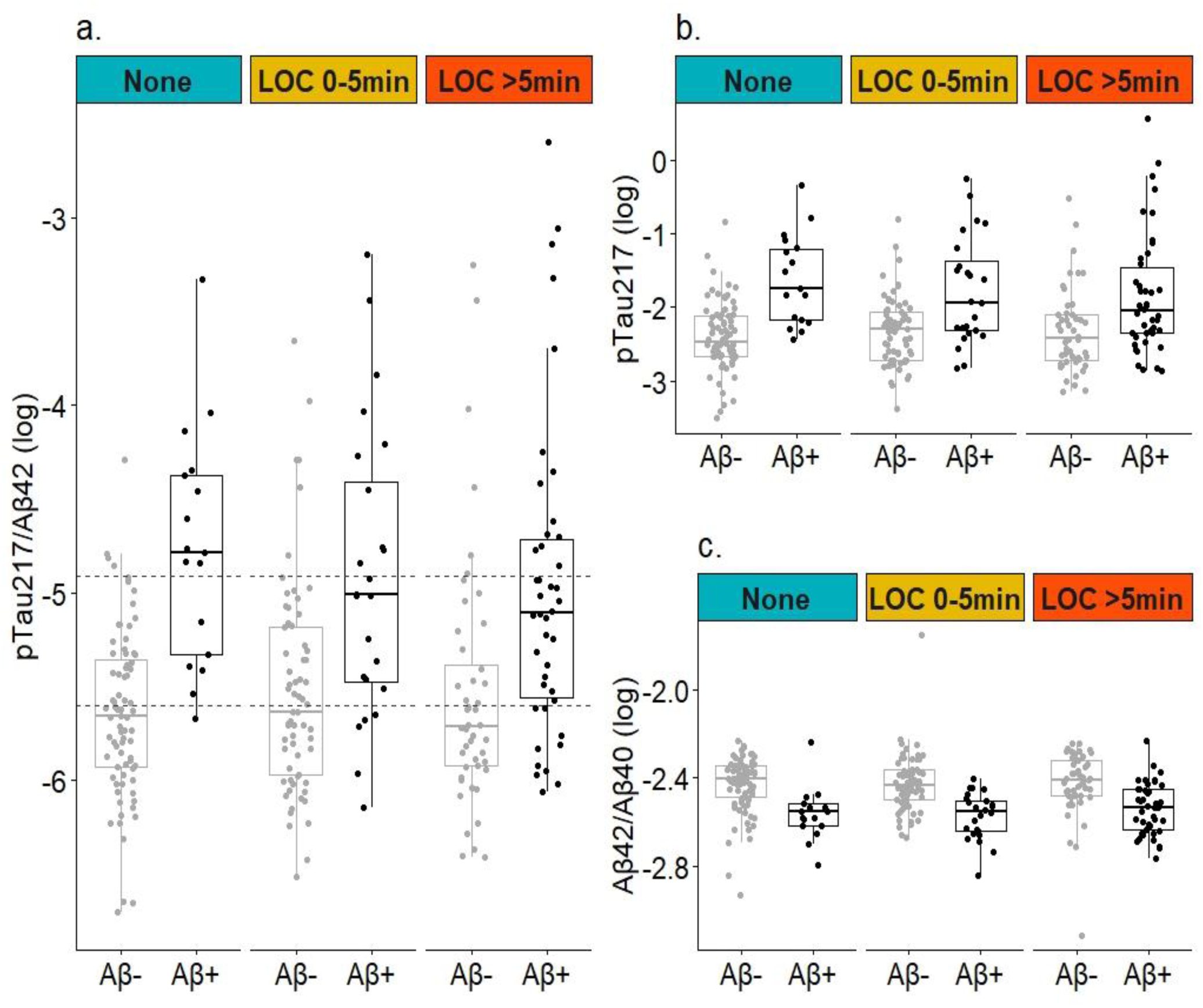
Biomarkers’ amyloid-β discrimination by Aβ-PET positivity (Aβ+/ Aβ-) and TBI group. Biomarker and biomarker ratios were log-transformed for figure clarity. a. Fujirebio Lumipulse pTau217/ Aβ42. Dashed lines indicate indeterminate zone (values above the upper dashed line are classified as Aβ+, values below the lower dashed line are classified as Aβ-). b. Fujirebio Lumipulse pTau217, c. Fujirebio Lumipulse Aβ42/ Aβ40. Abbreviations: Aβ= amyloid beta, LOC=Loss of consciousness, min=minutes, PET-positron emission tomography.

**Table 2.**
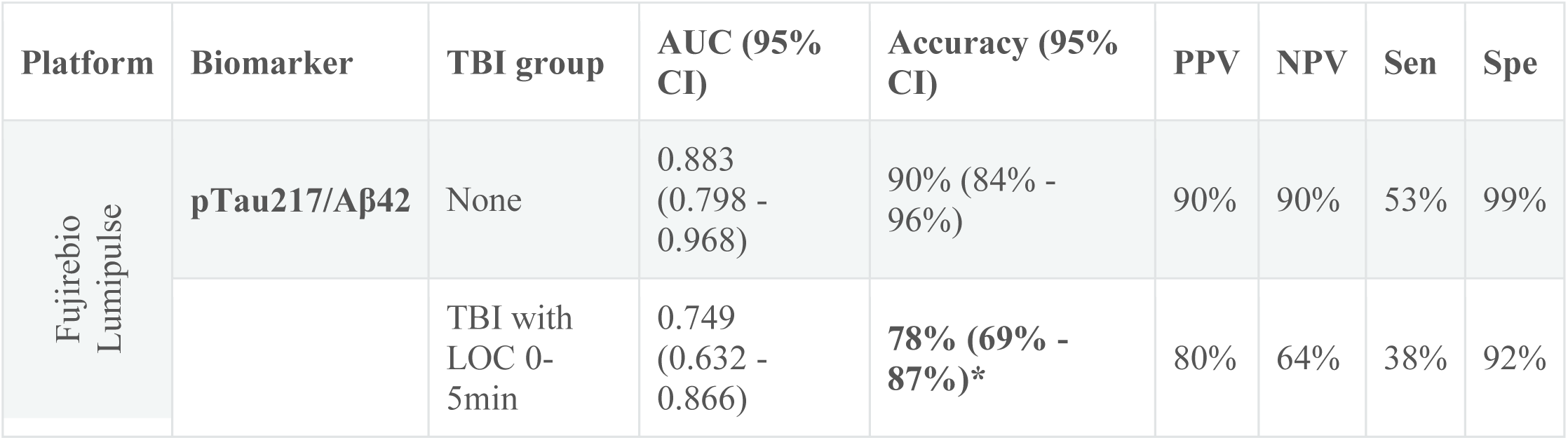
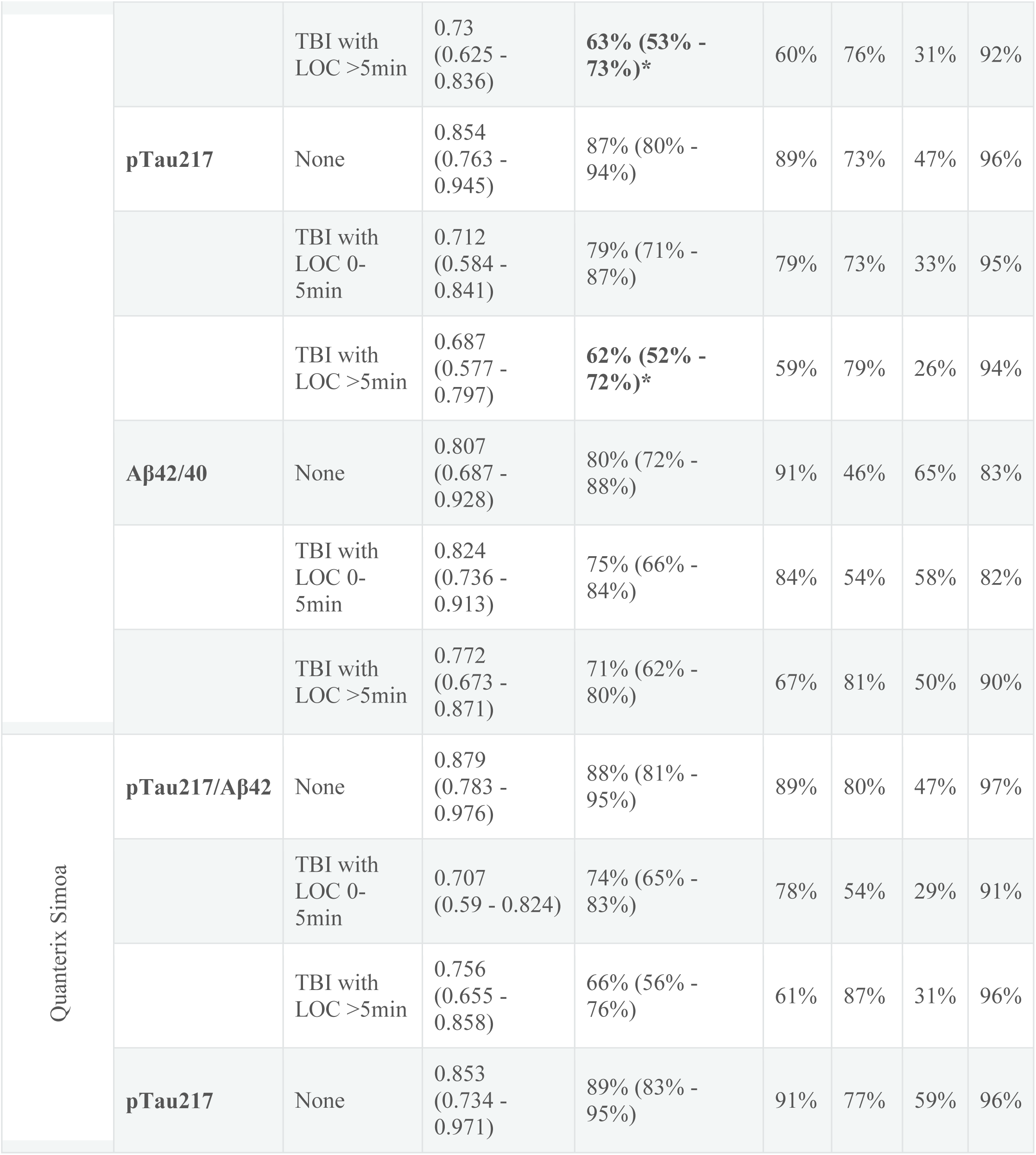
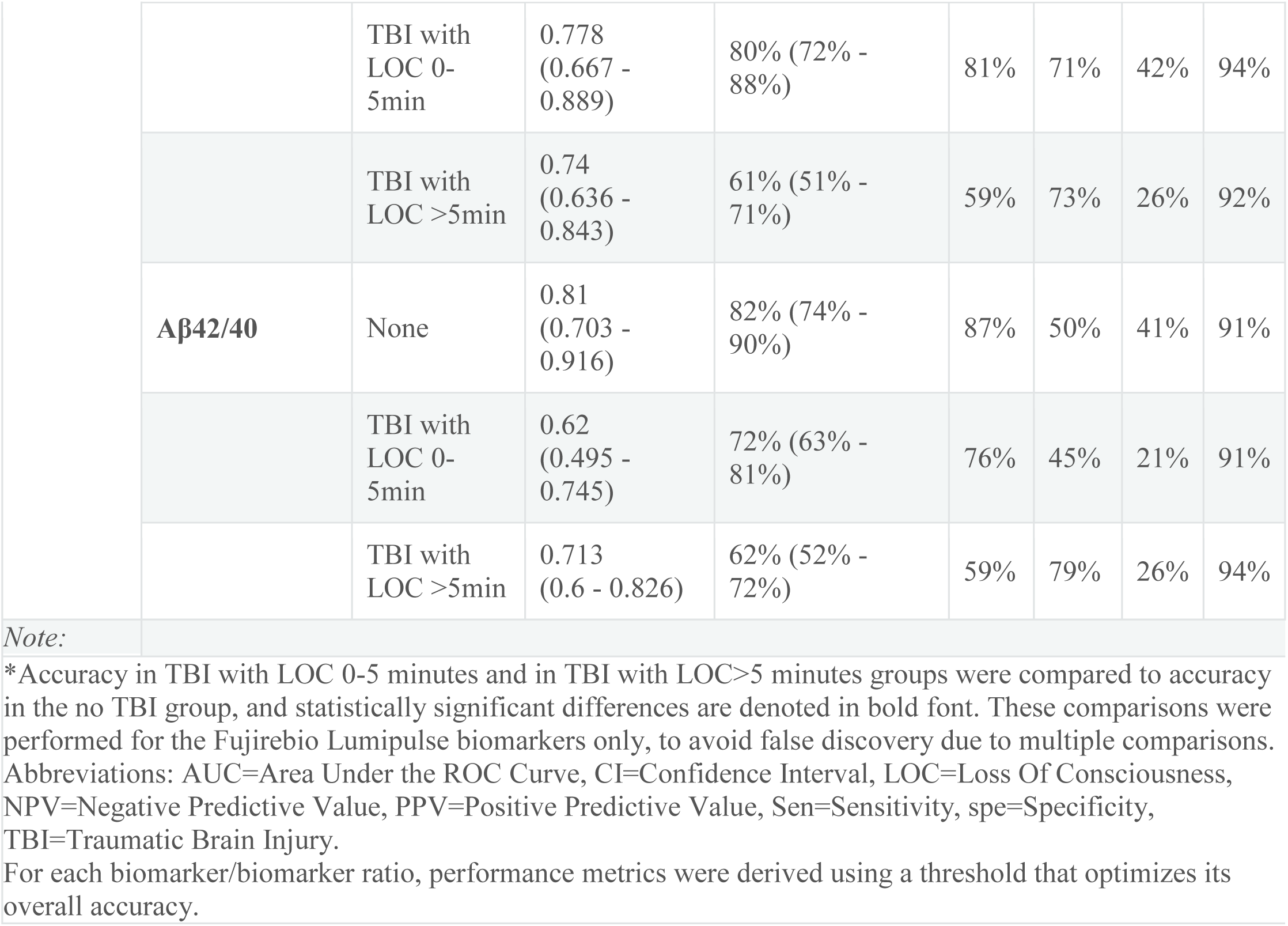
Aβ40/42, pTau217and pTau217/Aβ42 performance by TBI category.

Results were similar when applying the dual cut-off for Lumipulse pTau217/Aβ42 ratio (**Table 3**). Notably, 36.0% (N=98) of participants had to be excluded due to indeterminate results (33.7% of no TBI, 35.7% of TBI LOC 0-5 minutes, 30.6% of TBI LOC>5 minutes). Characteristics by Lumipulse result category are presented in **Supplementary Table S2**. Among Veterans with a positive or negative result (n=174), accuracy was 88% in those without TBI, 76% in those with TBI with LOC 0-5 minutes (*P*=0.093), and 62% in those with TBI with LOC>5 minutes (*P*<0.001).

**Table 3.**
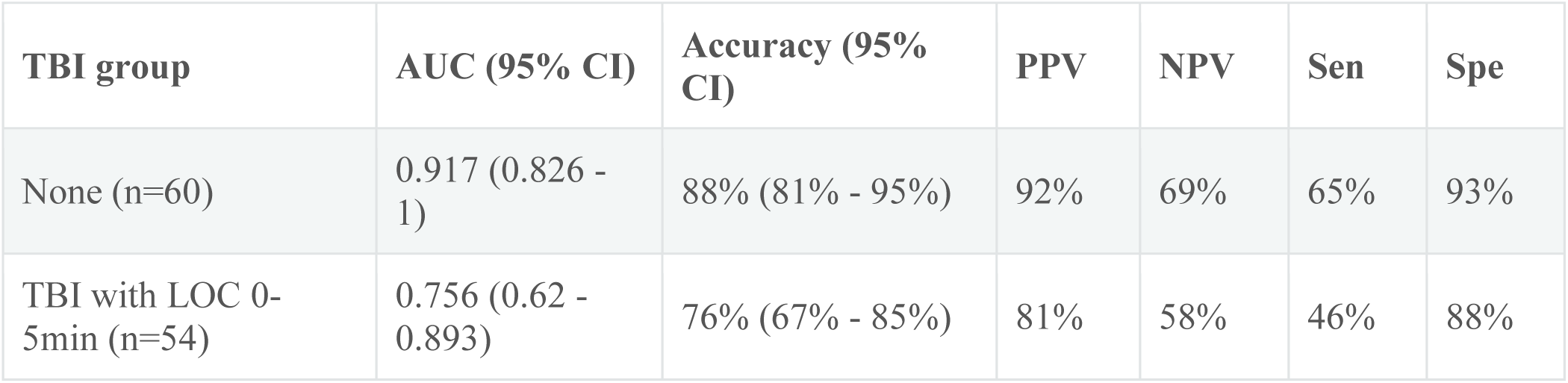
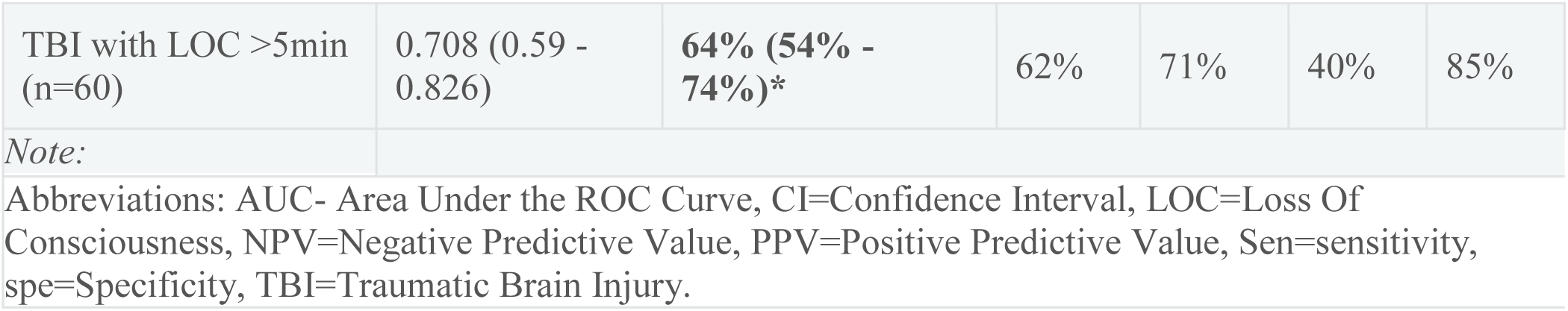
P-Tau217/Aβ 42 by TBI with dual cut-off.

| **TBI group** | **AUC (95% CI)** | **Accuracy (95% CI)** | **PPV** | **NPV** | **Sen** | **Spe** |
| --- | --- | --- | --- | --- | --- | --- |
| None (n=60) | 0.917 (0.826 - 1) | 88% (81% - 95%) | 92% | 69% | 65% | 93% |
| TBI with LOC 0-5min (n=54) | 0.756 (0.62 - 0.893) | 76% (67% - 85%) | 81% | 58% | 46% | 88% |
| TBI with LOC >5min<br>(n=60) | 0.708 (0.59 -<br>0.826) | **64% (54% -**<br>**74%)\*** | 62% | 71% | 40% | 85% |

*Note:*
Abbreviations: AUC- Area Under the ROC Curve, CI=Confidence Interval, LOC=Loss Of Consciousness, NPV=Negative Predictive Value, PPV=Positive Predictive Value, Sen=sensitivity, spe=Specificity, TBI=Traumatic Brain Injury.

Based on centiloids, less of half of Aβ-PETv+ Veterans with TBI would be identified as positive for Aβ (CL>40), compared to more than 75% of Aβ-PETv+ Veterans without TBI (Figure 2). Among Veterans without TBI, plasma NfL and GFAP were significantly higher among Aβ-PETv+ Veterans versus Aβ-PETv- Veterans (*P*=0.026, *P*<0.001, **Figure 2**). This association between Aβ-positivity and plasma NfL and GFAP level was not seen in Veterans with TBI (all *P*>0.08) in whom NfL and GFAP levels were similar regardless of Aβ-PETv status. Summaries of all biomarker and biomarker ratio levels by Aβ-PETv positivity and TBI group are presented in **supplementary Table S3**.

**Figure 2:**
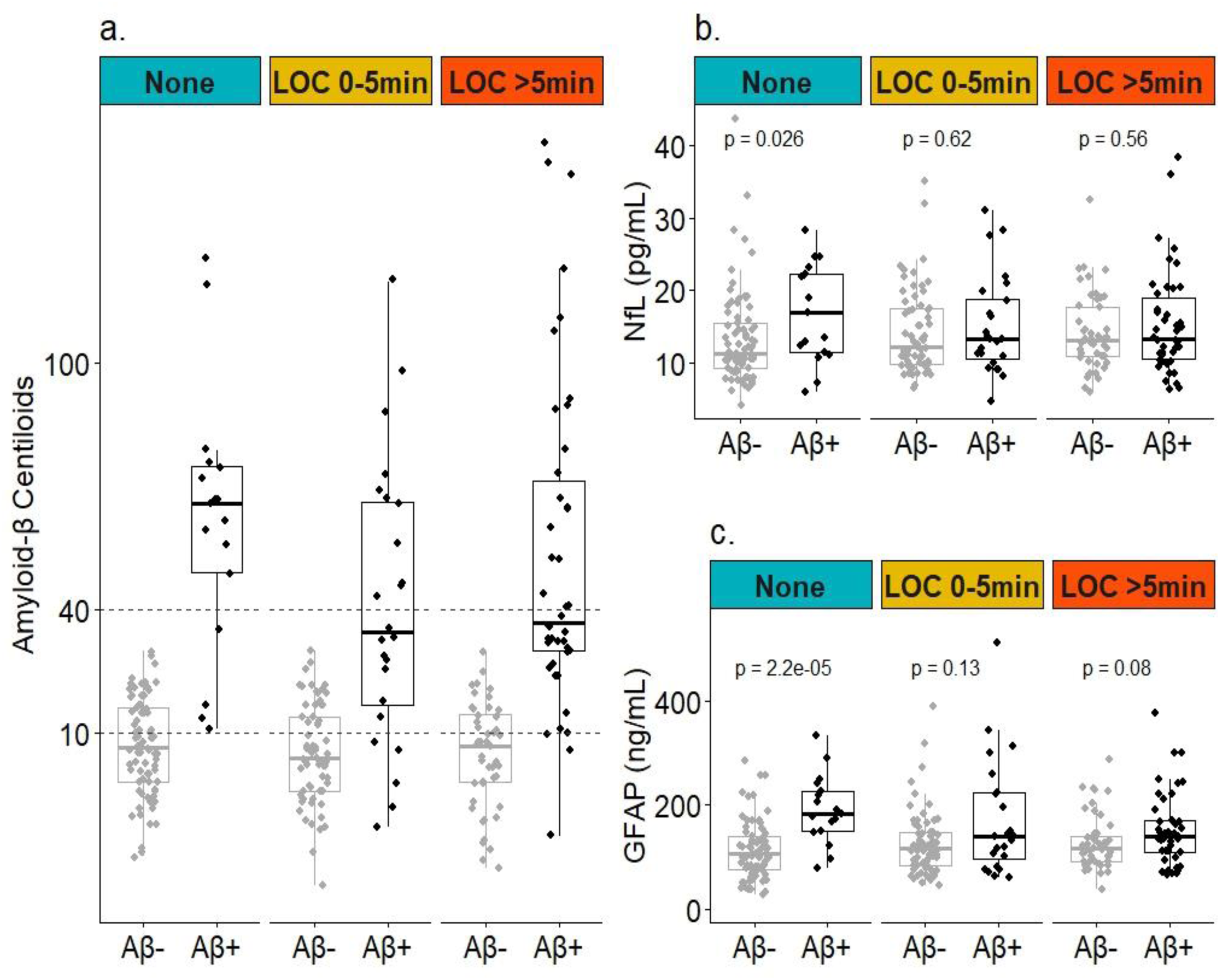
Distribution of Aβ centiloids , NfL and GFAP by Aβ-PET positivity (Aβ+/ Aβ-) and TBI group. a. Aβ centiloids, dashed lines indicate indeterminate zone (values above the upper dashed line are classified as Aβ+, values below the lower dashed line are classified as Aβ-), b. Neurofilament light chain (NfL), c. Glial Fibrillary Acidic Protein (GFAP).c.. *P*-values indicate comparisons performed using Wilcoxon-Mann-Whitney tests. Abbreviations: Aβ= amyloid beta, LOC=Loss of consciousness, min=minutes.

Overall findings were similar in our three sensitivity analyses, including analysis in Veterans who did not have a TBI in the past 10 years (no TBI n=93; TBI with LOC 0-5 minutes n=75; TBI with LOC>5 minutes n=78; **Supplementary Table S6**), comparison of results among cognitively unimpaired Veterans versus Veterans with MCI (**Supplementary Table S4**), and analysis using a quantitative SUVR threshold to define Aβ-PET positivity in lieu of consensus visual read (**Supplementary Table S5**).

## Discussion

We confirmed accuracy of 90% for the Lumipulse AD test in discrimination of Aβ-PET positivity among older Veterans with normal cognition or MCI with no TBI history. In older Veterans with prior TBI, the test’s accuracy using either a single cut-off (78%-63%) or the FDA-approved dual cut-off (82%-62%) did not meet the recommended threshold of 90% accuracy.^27^ Higher prevalence is often linked to higher PPV and sensitivity; however, despite higher Aβ-PETv+ prevalence, participants with TBI showed lower PPV and sensitivity. This was driven by a high proportion of false negatives (misdiagnosed Aβ-PETv+) and low proportion of true positives (correctly diagnosed Aβ-PETv+), meaning this subpopulation was underdiagnosed for AD, based on plasma biomarkers.

In Veterans without TBI history, Aβ-PETv+ was associated with significantly higher plasma GFAP and NfL levels, as expected. AD-associated elevation in plasma GFAP is believed to result from astrocytes surrounding Aβ plaques, even at an early stage of disease.^48^ In contrast, NfL is considered a later and non-specific marker of neurodegeneration that may be elevated in both AD and non-AD dementia as the disease progresses.^43^ Surprisingly, there was no association between Aβ-PETv+ and plasma GFAP or NfL levels among Veterans with TBI history, suggesting that brain amyloid is not the primary driver of glial activation or neurodegeneration in this cohort of mostly cognitively normal Veterans with TBI. Furthermore, among all Aβ-PETv+ Veterans (e.g., among Veterans with AD), those with TBI had a different distribution of plasma GFAP, plasma NfL and centiloids (indicating earlier stage of AD) compared to those without TBI. This finding suggests that Aβ pathophysiology may differ in Veterans with TBI..

The Lumipulse AD test dual cutoff yielded an indeterminate result for over a third of participants (33-39% across TBI groups), without improving accuracy or AUC. This substantial proportion of individuals who will require further evaluation is higher than the 10.6%–16.5% reported in a prior cohort of varying cognitive status.^49^ Research in cognitively unimpaired or mildly impaired individuals is needed to determine the percentage of indeterminate results in this target population and to understand why so many Veterans received one.

An incidental, but notable, finding was a significant association of TBI history with Aβ-PETv+, with a dose-response trend which contrasts with previous reports.^29^ Our analysis differs in TBI definition and categorization, and in using Aβ-PET visual reads instead of SUVR quantification for detecting Aβ positivity. Visual reads consider Aβ distribution patterns in addition to quantity, and are the current FDA-approved gold-standard method for determining Aβ-PET status for treatment purposes.^50,51^ Surprisingly, Aβ-PETv+ Veterans with TBI had lower centiloids compared to Aβ-PETv+ Veterans without TBI, highlighting the added value of visual PET rating and the complex relationship between TBI and Aβ.

Our cohort was mostly cognitively unimpaired (70%, and 30% with MCI). Previous studies of biomarkers’ ability to detect Aβ-PET+ in all or mostly cognitively unimpaired individuals had variable results. For plasma Aβ40/42 ratio, studies using the Enzyme-Linked ImmunoSorbent Assay (ELISA) and chemiluminescence enzyme immunoassay HISCL^®^ platforms found AUCs of 0.794^52^ and 0.968^53^, respectively. Another study using the Fujirebio Lumipulse platform found an AUC=0.81,^54^ same as our finding of AUC=0.807 for the no TBI group. For plasma pTau217, a meta-analysis of 30 studies found pooled sensitivity of 76% and specificity 83% in cognitively unimpaired (85% and 86% in cognitively impaired).^55^ We found sensitivity ranged between 41% (no TBI) and 26% (TBI with LOC>5min). A systematic review of biomarkers’ performance found that pTau217 had an AUC>0.9,^56^ and we found AUC ranged between 0.854 and 0.687. For plasma pTau217/Aβ42 ratio, a study using the Fujirebio Lumipulse platform found an AUC of 0.927,^57^ compared to our AUC of 0.879. In summary, while performance varies across platforms and cohorts, several prior studies align with our findings in Veterans without TBI. However, we found no prior reports showing performance as poor as that observed in Veterans with TBI.

Our findings of lower accuracy in persons with TBI history are supported by a similar study in persons with chronic traumatic encephalopathy (CTE), a neurodegenerative disease linked to chronic TBI and characterized by perivascular pTau deposition.^58^ This study^59^ found lower AUC for pTau217 in detection of Aβ-PET+ in persons with CTE (AUC=0.79), compared to persons with AD (AUC=0.86). In participants who were Aβ-PET+, persons with CTE had significantly lower plasma pTau217 compared to persons with no CTE. Recent evidence suggests that CTE is caused by TBI-triggered disruptions in waste clearance of pathogenic proteins, resulting in pTau deposition and neuroinflammation.^60^ Aβ deposition in the brain may not manifest in plasma pTau217 and/or Aβ42 due to co-pathology or altered clearance pathways,^61^ which may occur in persons with TBI history. This hypothesized mechanism is supported by similar findings for NfL and GFAP, which were lower in Aβ-PETv+ participants with TBI history compared to Aβ-PETv+ participants without TBI history. pTau217 has been shown to be AD-specific.^62,63^ For Aβ-PETv+ patients with a history of TBI, low levels of pTau217 may suggest that TBI rather than AD is the underlying or a contributing pathology. In such patients, biomarkers pTau217, Aβ42, GFAP, NfL, and Aβ-PET may prove useful to determine presence and stage of AD versus comorbid pathologies. Other factors to consider are chronic kidney disease, obesity, and cardiovascular conditions or medications that may affect blood AD biomarkers and may be more prevalent in Veterans with TBI.^8,11^

## Limitations

Our study has a few limitations. TBI history was assessed with a validated assessment tool, but data may have been affected by recall bias. Our cohort included almost only men. For better generalizability, the results of this study should be replicated in a sample enriched for TBI exposure in women and men, Veterans and civilians. Our cohort was enriched for PTSD, which was associated with TBI exposure. Larger studies will be powered to investigate this association and its implications on accuracy, as well as the role of age and APOEε4.

The pTau217/Aβ42 published performance metrics (92% PPV, 97% NVP) were derived from individuals exhibiting signs and symptoms of cognitive decline. Our cohort was mostly asymptomatic, however, the no TBI group had similar PPV (89%).

## Conclusions

Prior TBI modifies the accuracy of AD biomarkers, and care should be taken in interpreting AD blood test results in this context. Further research should validate these findings in larger diverse cohorts, including civilians with TBI, and establish guidelines for using biomarkers to determine Aβ-targeted therapy in persons with TBI history.

Investigating the mechanisms by which prior TBI confounds the AD biomarkers-Aβ-PET association is essential for optimizing precision diagnosis and treatment for both Veterans and civilians with TBI.

## Data Availability

Raw data is available on ADNI (https://adni.loni.usc.edu/). Data poduced in the present study is available upon request to the corresponding author.

## Acknowledgements

The authors would like to acknowledge the invaluable contribution of the Alzheimer’s Disease Neuroimaging Initiative (ADNI), especially the specific contributions of the ADNI DOD participants and investigators.

## Funding

This work was funded by the U.S. Department of Defense Congressionally Directed Medical Research Program (DoD CDMRP) Peer-Reviewed Alzheimer’s Research Program (PRARP; W81XWH-21-PRARP-ADRA to RCG) and U.S. National Institute of Health (NIH) National Institute of Aging (NIA). LCS, NH, and DLS were also supported by grant P30AG066518. AV, DSM, GB and RL were supported by NIA grant P30AG062422.

## Conflict of interests

All authors have completed the ICMJE uniform disclosure form at www.icmje.org/coi_disclosure.pdf and declare: neither Fujirebio nor Quanterix were directly involved in the design, execution, analysis, or interpretation of the findings presented here. Throughout the conduct of this work, RCG received additional unrelated research support from the U.S. National Institutes of Health, the U.S. Department of Defense Congressionally Directed Medical Research Program, the Binational Science Foundation, the Sheba Research Authority, and Eli Lilly and Co (in the form of donated doses of Florbetapir); RCG has an active academic collaboration with GryphonBio Inc. and an active non-disclosure agreement with C2N. No other relationships or activities that could appear to have influenced the submitted work.

## Data availability

Raw data is available on ADNI (https://adni.loni.usc.edu/). Analysis-ready data is available on request from the corresponding author.

## Clinical Trials Registration

This retrospective study that leveraged existing de-identified data and specimens that were prospectively collected for a different purpose is not listed on ClinicalTrials.gov.

## Notes

### Author Declarations

IRB of the Sheba Medical Center gave ethical approval for this work

## References

1. Zhong X, Wang Q, Yang M, et al. Plasma p-tau217 and p-tau217/Aβ1-42 are effective biomarkers for identifying CSF- and PET imaging-diagnosed Alzheimer’s disease: Insights for research and clinical practice. Alzheimer’s & Dementia. 2025;21(2):e14536. doi:10.1002/alz.14536

2. Barthélemy NR, Salvadó G, Schindler SE, et al. Highly accurate blood test for Alzheimer’s disease is similar or superior to clinical cerebrospinal fluid tests. Nat Med. 2024;30(4):1085–1095. doi:10.1038/s41591-024-02869-z

3. Rudolph MD, Sutphen CL, Register TC, et al. Evaluation of plasma p-tau217 for detecting amyloid pathology in a heterogeneous community-based cohort. Alzheimer’s & Dementia. 2025;21(7):e70426. doi:10.1002/alz.70426

4. Panza F, Lozupone M, Logroscino G, Imbimbo BP. A critical appraisal of amyloid-β-targeting therapies for Alzheimer disease. Nat Rev Neurol. 2019;15(2):73–88. doi:10.1038/s41582-018-0116-6

5. Karran E, De Strooper B. The amyloid hypothesis in Alzheimer disease: new insights from new therapeutics. Nat Rev Drug Discov. 2022;21(4):306–318. doi:10.1038/s41573-022-00391-w

6. Schindler SE, Petersen KK, Saef B, et al. Head-to-head comparison of leading blood tests for Alzheimer’s disease pathology. Alzheimer’s & Dementia. 2024;20(11):8074–8096. doi:10.1002/alz.14315

7. Cousins KAQ, Korecka M, Wan Y, et al. Comparison of plasma p-tau217/Aβ42, p-tau217, and Aβ42/Aβ40 biomarkers by race to detect Alzheimer’s disease. Alzheimer’s & Dementia. 2025;21(8):e70469. doi:10.1002/alz.70469

8. Mielke MM, Fowler NR. Alzheimer disease blood biomarkers: considerations for population-level use. Nat Rev Neurol. 2024;20(8):495–504. doi:10.1038/s41582-024-00989-1

9. Vespa J. Aging Veterans: America’s Veteran Population in Later Life. Published online July 2023. chrome-extension://efaidnbmnnnibpcajpcglclefindmkaj/https://www.census.gov/content/dam/Census/library/publications/2023/acs/acs-54.pdf

10. Raza Z, Hussain SF, Ftouni S, et al. Dementia in military and veteran populations: a review of risk factors—traumatic brain injury, post-traumatic stress disorder, deployment, and sleep. Military Med Res. 2021;8(1):1–13. doi:10.1186/s40779-021-00346-z

11. Gardner RC, Barnes DE, Li Y, Boscardin J, Peltz C, Yaffe K. Medical and Psychiatric Risk Factors for Dementia in Veterans with and without Traumatic Brain Injury (TBI): A Nationwide Cohort Study. J Prev Alzheimers Dis. 2023;10(2):244–250. doi:10.14283/jpad.2023.16

12. Kornblith ES, Yaffe K, Langa KM, Gardner RC. Prevalence of Lifetime History of Traumatic Brain Injury among Older Male Veterans Compared with Civilians: A Nationally Representative Study. Journal of Neurotrauma. Published online December 3, 2020. doi:10.1089/neu.2020.7062

13. Hoge CW, McGurk D, Thomas JL, Cox AL, Engel CC, Castro CA. Mild traumatic brain injury in U.S. Soldiers returning from Iraq. The New England journal of medicine. 2008;358(5):453–463. doi:10.1056/NEJMoa072972

14. Livingston G, Huntley J, Sommerlad A, et al. Dementia prevention, intervention, and care: 2020 report of the Lancet Commission. Lancet. 2020;396(10248):413–446. doi:10.1016/S0140-6736(20)30367-6

15. Barnes DE, Byers AL, Gardner RC, Seal KH, Boscardin WJ, Yaffe K. Association of Mild Traumatic Brain Injury With and Without Loss of Consciousness With Dementia in US Military Veterans. JAMA Neurol. 2018;75(9):1055–1061. doi:10.1001/jamaneurol.2018.0815

16. Gatson JW, Stebbins C, Mathews D, et al. Evidence of increased brain amyloid in severe TBI survivors at 1, 12, and 24 months after injury: report of 2 cases. Published online June 1, 2016. doi:10.3171/2015.6.JNS15639

17. Gardner RC, Possin KL, Hess CP, et al. Evaluating and treating neurobehavioral symptoms in professional American football players: Lessons from a case series. Neurology Clinical practice. 2015;5(4):285–295. doi:10.1212/CPJ.0000000000000157

18. Hossain I, Marklund N, Czeiter E, Hutchinson P, Buki A. Blood biomarkers for traumatic brain injury: A narrative review of current evidence. Brain Spine. 2023;4:102735. doi:10.1016/j.bas.2023.102735

19. Gill J, Latour L, Diaz-Arrastia R, et al. Glial fibrillary acidic protein elevations relate to neuroimaging abnormalities after mild TBI. Neurology. 2018;91(15):e1385–e1389. doi:10.1212/WNL.0000000000006321

20. Gardner RC, Rubenstein R, Wang KKW, et al. Age-Related Differences in Diagnostic Accuracy of Plasma Glial Fibrillary Acidic Protein and Tau for Identifying Acute Intracranial Trauma on Computed Tomography: A TRACK-TBI Study. Journal of Neurotrauma. 2018;35(20):2341–2350. doi:10.1089/neu.2018.5694

21. Rubenstein R, Chang B, Yue JK, et al. Comparing Plasma Phospho Tau, Total Tau, and Phospho Tau–Total Tau Ratio as Acute and Chronic Traumatic Brain Injury Biomarkers. JAMA Neurol. 2017;74(9):1063–1072. doi:10.1001/jamaneurol.2017.0655

22. Rubenstein R, McQuillan L, Wang KKW, et al. Temporal Profiles of P-Tau, T-Tau, and P-Tau:Tau Ratios in Cerebrospinal Fluid and Blood from Moderate-Severe Traumatic Brain Injury Patients and Relationship to 6–12 Month Global Outcomes. Journal of Neurotrauma. Published online January 31, 2024. doi:10.1089/neu.2022.0479

23. Bogoslovsky Tanya, Wilson David, Chen Yao, et al. Increases of Plasma Levels of Glial Fibrillary Acidic Protein, Tau, and Amyloid β up to 90 Days after Traumatic Brain Injury. Journal of Neurotrauma. Published online January 1, 2017. doi:10.1089/neu.2015.4333

24. Whiteneck GG, Cuthbert JP, Corrigan JD, Bogner JA. Prevalence of Self-Reported Lifetime History of Traumatic Brain Injury and Associated Disability: A Statewide Population-Based Survey. The Journal of Head Trauma Rehabilitation. 2016;31(1):E55. doi:10.1097/HTR.0000000000000140

25. Karamian A, Lucke-Wold B, Seifi A. Prevalence of Traumatic Brain Injury in the General Adult Population of the USA: A Meta-Analysis. Neuroepidemiology. Published online August 22, 2024:1–10. doi:10.1159/000540676

26. Gardner RC, Rivera E, O’Grady M, et al. Screening for Lifetime History of Traumatic Brain Injury Among Older American and Irish Adults at Risk for Dementia: Development and Validation of a Web-Based Survey. J Alzheimers Dis. 2020;74(2):699–711. doi:10.3233/JAD-191138

27. Jack CR, Andrews JS, Beach TG, et al. Revised criteria for diagnosis and staging of Alzheimer’s disease: Alzheimer’s Association Workgroup. doi:10.1002/alz.13859

28. Weiner MW, Harvey D, Hayes J, et al. Effects of traumatic brain injury and posttraumatic stress disorder on development of Alzheimer’s disease in Vietnam Veterans using the Alzheimer’s Disease Neuroimaging Initiative: Preliminary Report. Alzheimers Dement (N Y). 2017;3(2):177–188. doi:10.1016/j.trci.2017.02.005

29. Weiner MW, Harvey D, Landau SM, et al. Traumatic brain injury and post-traumatic stress disorder are not associated with Alzheimer’s disease pathology measured with biomarkers. Alzheimer’s & Dementia. 2023;19(3):884–895. doi:10.1002/alz.12712

30. Centiloid recommendations for clinical context-of-use from the AMYPAD consortium - Collij - 2024 - Alzheimer’s & Dementia - Wiley Online Library. Accessed August 4, 2025. https://alz-journals-onlinelibrary-wiley-com.sheba.portium.org/doi/full/10.1002/alz.14336?

31. Landau SM, Ward TJ, Murphy A, et al. Quantification of amyloid beta and tau PET without a structural MRI. Alzheimer’s & Dementia. 2023;19(2):444–455. 10.1002/alz.12668

32. Corrigan JD, Bogner J. Initial reliability and validity of the Ohio State University TBI Identification Method. J Head Trauma Rehabil. 2007;22(6):318–329. doi:10.1097/01.HTR.0000300227.67748.77

33. Bogner J, Corrigan JD. Reliability and predictive validity of the Ohio State University TBI identification method with prisoners. J Head Trauma Rehabil. 2009;24(4):279–291. doi:10.1097/HTR.0b013e3181a66356

34. Bogner J, Corrigan JD, Yi H, et al. Lifetime History of Traumatic Brain Injury and Behavioral Health Problems in a Population-Based Sample. The Journal of Head Trauma Rehabilitation. 2020;35(1):E43. doi:10.1097/HTR.0000000000000488

35. McKinlay A, Corrigan JD, Bogner JA, Horwood LJ. Obtaining a History of Childhood Traumatic Brain Injury Using the Ohio State University TBI Identification Method to Elicit Adult Recall. The Journal of Head Trauma Rehabilitation. 2017;32(6):E24. doi:10.1097/HTR.0000000000000284

36. Manley GT, Dams-O’Connor K, Alosco ML, et al. A new characterisation of acute traumatic brain injury: the NIH-NINDS TBI Classification and Nomenclature Initiative. The Lancet Neurology. 2025;24(6):512–523. doi:10.1016/S1474-4422(25)00154-1

37. Silverberg ND, Iverson GL, Cogan A, et al. The American Congress of Rehabilitation Medicine Diagnostic Criteria for Mild Traumatic Brain Injury. Archives of Physical Medicine and Rehabilitation. 2023;104(8):1343–1355. doi:10.1016/j.apmr.2023.03.036

38. Peltz CB, Gardner RC, Kenney K, Diaz-Arrastia R, Kramer JH, Yaffe K. Neurobehavioral Characteristics of Older Veterans With Remote Traumatic Brain Injury. J Head Trauma Rehabil. 2017;32(1):E8–E15. doi:10.1097/HTR.0000000000000245

39. Gardner RC, Peltz CB, Kenney K, Covinsky KE, Diaz-Arrastia R, Yaffe K. Remote Traumatic Brain Injury Is Associated with Motor Dysfunction in Older Military Veterans. J Gerontol A Biol Sci Med Sci. 2017;72(9):1233–1238. doi:10.1093/gerona/glw341

40. Alcolea D, Beeri MS, Rojas JC, Gardner RC, Lleó A. Blood Biomarkers in Neurodegenerative Diseases. Neurology. 2023;101(4):172–180. doi:10.1212/WNL.0000000000207193

41. Yang Z, Wang KKW. Glial fibrillary acidic protein: from intermediate filament assembly and gliosis to neurobiomarker. Trends in Neurosciences. 2015;38(6):364–374. doi:10.1016/j.tins.2015.04.003

42. Jung Y, Damoiseaux JS. The potential of blood neurofilament light as a marker of neurodegeneration for Alzheimer’s disease. Brain. 2024;147(1):12–25. doi:10.1093/brain/awad267

43. Gaetani L, Blennow K, Calabresi P, Filippo MD, Parnetti L, Zetterberg H. Neurofilament light chain as a biomarker in neurological disorders. J Neurol Neurosurg Psychiatry. 2019;90(8):870–881. doi:10.1136/jnnp-2018-320106

44. Mattsson N, Andreasson U, Zetterberg H, Blennow K, for the Alzheimer’s Disease Neuroimaging Initiative. Association of Plasma Neurofilament Light With Neurodegeneration in Patients With Alzheimer Disease. JAMA Neurol. 2017;74(5):557–566. doi:10.1001/jamaneurol.2016.6117

45. Cicognola C, Janelidze S, Hertze J, et al. Plasma glial fibrillary acidic protein detects Alzheimer pathology and predicts future conversion to Alzheimer dementia in patients with mild cognitive impairment. Alz Res Therapy. 2021;13(1):68. doi:10.1186/s13195-021-00804-9

46. Korecka M. ADNI3 Biomarker Biofluid Collection, Processing and Shipment. https://adni.loni.usc.edu/wp-content/themes/freshnews-dev-v2/documents/bio/ADNI3_Biomarker_Sample_Collection_Processing_and_Shipment.pdf

47. R Core Team (2022). R: A language and environment for statistical computing. R Foundation for Statistical Computing, Vienna, Austria. URL https://www.R-project.org/.

48. Fernández-Matarrubia M, Valera-Barrero A, Renuncio-García M, et al. Early microglial and astrocyte reactivity in preclinical Alzheimer’s disease. Alzheimers Dement. 2025;21(8):e70502. doi:10.1002/alz.70502

49. Wang J, Huang S, Lan G, et al. Diagnostic accuracy of plasma p-tau217/Aβ42 for Alzheimer’s disease in clinical and community cohorts. doi:10.1002/alz.70038

50. Zeltzer E, Schonhaut DR, Mundada NS, et al. Concordance between amyloid-PET quantification and real-world visual reads: results from IDEAS. medRxiv. Preprint posted online November 4, 2024:2024.10.31.24316518. doi:10.1101/2024.10.31.24316518

51. de Bruin H, Groot C, Kamps S, et al. Amyloid-β and tau deposition in traumatic brain injury: a study of Vietnam War veterans. Brain Commun. 2025;7(1):fcaf009. doi:10.1093/braincomms/fcaf009

52. Vergallo A, Mégret L, Lista S, et al. Plasma amyloid β 40/42 ratio predicts cerebral amyloidosis in cognitively normal individuals at risk for Alzheimer’s disease. Alzheimer’s & Dementia. 2019;15(6):764–775. doi:10.1016/j.jalz.2019.03.009

53. Kubota M, Bun S, Takahata K, et al. Plasma biomarkers for early detection of alzheimer’s disease: a cross-sectional study in a Japanese cohort. Alz Res Therapy. 2025;17(1):131. doi:10.1186/s13195-025-01778-8

54. Figdore DJ, Wiste HJ, Bornhorst JA, et al. Performance of the Lumipulse plasma Aβ42/40 and pTau181 immunoassays in the detection of amyloid pathology. *Alzheimer’s & Dementia: Diagnosis*, Assessment & Disease Monitoring. 2024;16(1):e12545. doi:10.1002/dad2.12545

55. Khalafi M, Dartora WJ, McIntire LBJ, et al. Diagnostic accuracy of phosphorylated tau217 in detecting Alzheimer’s disease pathology among cognitively impaired and unimpaired: A systematic review and meta-analysis. doi:10.1002/alz.14458

56. Suresh S, Maffei L, Bauermeister S, Raymont V. Blood Biomarkers for Diagnosis & Differential Diagnosis of Alzheimer’s Disease in Real-World Clinical Populations: A Systematic Review. medRxiv. Preprint posted online July 7, 2025:2025.07.06.25330840. doi:10.1101/2025.07.06.25330840

57. Lehmann S, Gabelle A, Duchiron M, et al. Comparative performance of plasma pTau181/Aβ42, pTau217/Aβ42 ratios, and individual measurements in detecting brain amyloidosis. eBioMedicine. 2025;117. doi:10.1016/j.ebiom.2025.105805

58. de Sena Barbosa MG, Francisco GG de OA, de Souza RLV, et al. Chronic traumatic encephalopathy in athletes, players, boxers and military: systematic review. Annals of Medicine and Surgery. 2024;86(12):7238. doi:10.1097/MS9.0000000000002693

59. Asken BM, Tanner JA, VandeVrede L, et al. Plasma P-tau181 and P-tau217 in Patients With Traumatic Encephalopathy Syndrome With and Without Evidence of Alzheimer Disease Pathology. Neurology. 2022;99(6):e594–e604. doi:10.1212/WNL.0000000000200678

60. Barker RB, Karakaya E, Baran D, et al. The glymphatic and meningeal lymphatic systems may converge, connecting traumatic brain injury progression with chronic traumatic encephalopathy onset. Molecular and Cellular Neuroscience. 2025;134:104031. doi:10.1016/j.mcn.2025.104031

61. Ullah R, Lee EJ. Advances in Amyloid-β Clearance in the Brain and Periphery: Implications for Neurodegenerative Diseases. Experimental Neurobiology. 2023;32(4):216–246. doi:10.5607/en23014

62. Palmqvist S, Janelidze S, Quiroz YT, et al. Discriminative Accuracy of Plasma Phospho-tau217 for Alzheimer Disease vs Other Neurodegenerative Disorders. JAMA. 2020;324(8):772–781. doi:10.1001/jama.2020.12134

63. Thijssen EH, Joie RL, Strom A, et al. Plasma phosphorylated tau 217 and phosphorylated tau 181 as biomarkers in Alzheimer’s disease and frontotemporal lobar degeneration: a retrospective diagnostic performance study. The Lancet Neurology. 2021;20(9):739–752. doi:10.1016/S1474-4422(21)00214-3

64. Van Calster B, Nieboer D, Vergouwe Y, De Cock B, Pencina MJ, Steyerberg EW. A calibration hierarchy for risk models was defined: from utopia to empirical data. J Clin Epidemiol. 2016;74:167–176. doi:10.1016/j.jclinepi.2015.12.005

