## Supplements for "Effect of prior traumatic brain injury on Alzheimer’s disease blood biomarkers in Vietnam Veterans"

### Supplementary Table S1. Traumatic Brain Injury characteristics

| N (%) | **Overall**<br>N = 272[1] | **None**<br>N = 93[1] | **TBI with LOC 0-5min**<br>N = 89[1] | **TBI with LOC >5min**<br>N = 90[1] |
|---|---|---|---|---|
| **Worst TBI severity[2]** | | | | |
| No TBI | 93 (34%) | 93 (100%) | - | - |
| No LOC | 51 (19%) | - | 51 (57%) | - |
| LOC<30min | 84 (31%) | - | 38 (43%) | 46 (51%) |
| LOC>30min | 44 (16%) | - | - | 44 (49%) |
| **Longest LOC duration** | | | | |
| No LOC | 144 (53%) | 93 (100%) | 51 (57%) | - |
| 0-5 minutes | 38 (14%) | - | 38 (43%) | - |
| 5-9 minutes | 19 (7.0%) | - | - | 19 (21%) |
| 10-29 minutes | 27 (9.9%) | - | - | 27 (30%) |
| 30 minutes-24 hours | 30 (11%) | - | - | 30 (33%) |
| >24 hours | 14 (5.1%) | - | - | 14 (16%) |
| **Longest AOC duration** | | | | |
| No AOC | 120 (44%) | 93 (100%) | 13 (15%) | 14 (16%) |
| 0-5 minutes | 26 (9.6%) | - | 22 (25%) | 4 (4.4%) |
| 5-9 minutes | 9 (3.3%) | - | 6 (6.7%) | 3 (3.3%) |
| 10-29 minutes | 13 (4.8%) | - | 4 (4.5%) | 9 (10%) |
| 30 minutes-24 hours | 38 (14%) | - | 14 (16%) | 24 (27%) |
| >24 hours | 66 (24%) | - | 30 (34%) | 36 (40%) |
| **Longest PTA duration** | | | | |
| No PTA | 218 (80%) | 93 (100%) | 68 (76%) | 57 (63%) |
| 0-5 minutes | 4 (1.5%) | - | 1 (1.1%) | 3 (3.3%) |
| 5-9 minutes | 2 (0.7%) | - | 0 (0%)- | 2 (2.2%) |
| 10-29 minutes | 4 (1.5%) | - | 2 (2.2%) | 2 (2.2%) |
| 30 minutes-24 hours | 12 (4.4%) | - | 4 (4.5%) | 8 (8.9%) |
| >24 hours | 32 (12%) | - | 14 (16%) | 18 (20%) |
| **Number of TBIs with LOC** | | | | |
| 0 | 139 (51%) | 93 (100%) | 46 (52%) | - |
| 1 | 94 (35%) | - | 29 (33%) | 65 (72%) |
| 2 | 27 (9.9%) | - | 12 (13%) | 15 (17%) |
| 3 | 8 (2.9%) | - | 2 (2.2%) | 6 (6.7%) |
| 4 | 2 (0.7%) | - | 0 (0%) | 2 (2.2%) |
| 5 | 2 (0.7%) | - | 0 (0%) | 2 (2.2%) |

| | | | | |
|---|---|---|---|---|
| **TBI frequency** | | | | |
| 0 | 93 (34%) | 93 (100%) | - | - |
| 1 | 108 (40%) | - | 60 (67%) | 48 (53%) |
| 2 | 47 (17%) | - | 22 (25%) | 25 (28%) |
| 3 | 14 (5.1%) | - | 5 (5.6%) | 9 (10%) |
| 4 | 5 (1.8%) | - | 1 (1.1%) | 4 (4.4%) |
| 5 | 4 (1.5%) | - | 1 (1.1%) | 3 (3.3%) |
| 7 | 1 (0.4%) | - | 0 (0%) | 1 (1.1%) |
| **TBI severity and frequency** | | | | |
| No TBI | 93 (38%) | 93 (100%) | - | - |
| Single mild TBI without LOC | 40 (16%) | - | 40 (56%) | - |
| single mild TBI with LOC | 44 (18%) | - | 20 (28%) | 24 (30%) |
| 2+ mild TBIs | 24 (9.8%) | - | 12 (17%) | 12 (15%) |
| Single moderate-severe TBI | 38 (16%) | - | 0 (0%) | 38 (48%) |
| 2+ moderate-severe TBIs | 6 (2.4%) | 0 (0%) | 0 (0%) | 6 (7.5%) |
| Unknown | 27 | 0 | 17 | 10 |

[1]n (%)

[2]Abbreviations: AOC = Alteration of consciousness, LOC = Loss Of Consciousness, PTA = Post-Traumatic Amnesia, TBI = Traumatic Brain Injury

### Supplementary Table S2. Demographic and baseline characteristics by AD classification using pTau217/Aβ42 dual thresholds

| Median (IQR), or N (%) | **Overall**<br>N = 272[1] | **Positive**<br>N = 51[1] | **Negative**<br>N = 123[1] | **Indeterminate**<br>N = 98[1] |
|---|---|---|---|---|
| **Age** | 68.9 (66.7, 72.1) | 70.2 (67.8, 75.0) | 68.2 (66.3, 71.5) | 69.1 (66.7, 72.0) |
| **Race** | | | | |
| White | 231 (85%) | 44 (86%) | 102 (83%) | 85 (87%) |
| Black | 20 (7.4%) | 4 (7.8%) | 10 (8.1%) | 6 (6.1%) |
| Other | 21 (7.7%) | 3 (5.9%) | 11 (8.9%) | 7 (7.1%) |
| **Years of education** | 15 (13, 17) | 15 (13, 18) | 14 (13, 16) | 16 (14, 17) |
| **APOE ε4 carrier** | 68 (26%) | 25 (51%) | 20 (17%) | 23 (25%) |
| (Missing) | 13 | 2 | 6 | 5 |
| **Cognitive status** | | | | |
| Normal | 227 (83%) | 42 (82%) | 104 (85%) | 81 (83%) |
| MCI | 45 (17%) | 9 (18%) | 19 (15%) | 17 (17%) |

| | | | | |
|---|---|---|---|---|
| **Baseline Clinical Dementia Rating** | | | | |
| 0 | 181 (70%) | 32 (67%) | 75 (65%) | 74 (77%) |
| 0.5 | 79 (30%) | 16 (33%) | 41 (35%) | 22 (23%) |
| (Missing) | 12 | 3 | 7 | 2 |
| **Baseline ECog-39** | 59 (48, 77) | 54 (47, 77) | 63 (48, 77) | 56 (48, 72) |
| **Aβ PET visual read** | | | | |
| Amyloid+ | 83 (31%) | 35 (69%) | 16 (13%) | 32 (33%) |
| Amyloid- | 189 (69%) | 16 (31%) | 107 (87%) | 66 (67%) |
| **Prior TBI in the past year** | | | | |
| TBI in the past year | 15 (5.5%) | 2 (3.9%) | 8 (6.5%) | 5 (5.1%) |
| TBI 1-10 years ago | 11 (4.0%) | 1 (2.0%) | 6 (4.9%) | 4 (4.1%) |
| TBI over 10 years ago | 153 (56%) | 33 (65%) | 64 (52%) | 56 (57%) |
| No TBI | 93 (34%) | 15 (29%) | 45 (37%) | 33 (34%) |
| **Years since prior TBI (where applicable)** | 46 (34, 49) | 47 (31, 49) | 46 (26, 49) | 47 (38, 50) |
| (Missing) | 93 | 15 | 45 | 33 |

[1]Median (Q1, Q3); n (%)

### Supplementary Table S3. Biomarker levels by TBI group and amyloid PET visual read

| | None | | | TBI with LOC 0-5min | | | TBI with LOC >5min | | |
|---|---|---|---|---|---|---|---|---|---|
| | **Aβ+** N = 17 | **Aβ-** N = 76 | **p-value**[1] | **Aβ+** N = 24 | **Aβ-** N = 65 | **p-value**[1] | **Aβ+** N = 42 | **Aβ-** N = 48 | **p-value**[2] |
| **Fujirebio Lumipulse pTau217/Aβ42** | | | **<0.001** | | | **<0.001** | | | **<0.001** |
| Median (Q1, Q3) | 0.008 (0.005, 0.013) | 0.003 (0.003, 0.005) | | 0.007 (0.004, 0.013) | 0.004 (0.003, 0.006) | | 0.006 (0.004, 0.009) | 0.003 (0.003, 0.005) | |
| (Min, Max) | (0.003, 0.036) | (0.001, 0.014) | | (0.002, 0.041) | (0.001, 0.026) | | (0.002, 0.075) | (0.002, 0.039) | |
| **Quanterix Simoa pTau217/Aβ42** | | | **<0.001** | | | **0.003** | | | **<0.001** |
| Median (Q1, Q3) | 0.08 (0.06, 0.14) | 0.03 (0.03, 0.05) | | 0.05 (0.04, 0.10) | 0.04 (0.03, 0.05) | | 0.06 (0.04, 0.10) | 0.03 (0.03, 0.05) | |

| | | | | | | | | | |
|---|---|---|---|---|---|---|---|---|---|
| (Min, Max) | (0.03, 0.22) | (0.01, 0.16) | | (0.03, 0.24) | (0.02, 0.31) | | (0.02, 0.31) | (0.02, 0.41) | |
| **Fujirebio Lumipulse pTau217 (pg/mL)** | | | **<0.001** | | | **0.002** | | | **0.002** |
| Median (Q1, Q3) | 0.18 (0.11, 0.30) | 0.09 (0.07, 0.12) | | 0.15 (0.10, 0.27) | 0.10 (0.07, 0.13) | | 0.13 (0.10, 0.25) | 0.09 (0.07, 0.12) | |
| (Min, Max) | (0.09, 0.71) | (0.03, 0.43) | | (0.06, 0.77) | (0.03, 0.45) | | (0.06, 1.76) | (0.04, 0.60) | |
| **Quanterix Simoa pTau217 (pg/mL)** | | | **<0.001** | | | **<0.001** | | | **<0.001** |
| Median (Q1, Q3) | 0.65 (0.44, 0.82) | 0.31 (0.25, 0.37) | | 0.51 (0.34, 0.87) | 0.32 (0.25, 0.38) | | 0.42 (0.34, 0.69) | 0.30 (0.24, 0.38) | |
| (Min, Max) | (0.21, 1.70) | (0.13, 2.20) | | (0.25, 1.40) | (0.16, 0.71) | | (0.19, 1.56) | (0.15, 1.17) | |
| **Fujirebio Lumipulse Aβ42/Aβ40** | | | **<0.001** | | | **<0.001** | | | **<0.001** |
| Median (Q1, Q3) | 0.078 (0.073, 0.081) | 0.090 (0.083, 0.096) | | 0.078 (0.071, 0.082) | 0.088 (0.082, 0.094) | | 0.079 (0.072, 0.086) | 0.090 (0.083, 0.099) | |
| (Min, Max) | (0.061, 0.107) | (0.053, 0.108) | | (0.058, 0.090) | (0.069, 0.173) | | (0.063, 0.107) | (0.044, 0.106) | |
| **Quanterix Simoa Aβ42/Aβ40** | | | **<0.001** | | | 0.085 | | | **<0.001** |
| Median (Q1, Q3) | 0.032 (0.029, 0.035) | 0.038 (0.035, 0.041) | | 0.035 (0.032, 0.038) | 0.037 (0.033, 0.041) | | 0.033 (0.031, 0.040) | 0.040 (0.036, 0.042) | |
| (Min, Max) | (0.018, 0.040) | (0.021, 0.051) | | (0.025, 0.059) | (0.015, 0.115) | | (0.023, 0.104) | (0.021, 0.099) | |
| **Quanterix Simoa GFAP (ng/mL)** | | | **<0.001** | | | 0.13 | | | 0.055 |
| Median (Q1, Q3) | 182 (150, 226) | 106 (75, 141) | | 140 (91, 223) | 116 (83, 146) | | 142 (110, 190) | 118 (92, 140) | |
| (Min, Max) | (77, 333) | (27, 284) | | (59, 511) | (45, 390) | | (65, 745) | (37, 288) | |
| **Quanterix Simoa NfL (pg/mL)** | | | **0.026** | | | 0.6 | | | 0.6 |
| Median (Q1, Q3) | 16.9 (11.5, 22.3) | 11.3 (9.2, 15.7) | | 13.3 (10.3, 19.2) | 12.2 (9.8, 17.5) | | 13.3 (10.3, 19.5) | 13.1 (10.8, 17.8) | |

| | | | | | | | | |
|---|---|---|---|---|---|---|---|---|
| (Min, Max) | (5.9, 28.2) | (4.2, 43.7) | | (4.6, 31.1) | (6.4, 35.1) | | (6.3, 38.4) | (5.8, 32.5) |

[1]Wilcoxon rank sum test

[2]Wilcoxon rank sum test; Wilcoxon rank sum exact test

### Supplementary Table S4. Aβ40/42, pTau217and pTau217/Aβ42 performance by cognitive status (Normal/MCI) vs amyloid PET visual read

*Supplementary Table S4. Aβ40/42, pTau217and pTau217/Aβ42 performance by cognitive status*

| Platform | Biomarker | Cognitive status | AUC (95% CI) | Accuracy (95% CI) | PPV | NPV | Sen | Spe |
|---|---|---|---|---|---|---|---|---|
| Fujirebio Lumipulse | **pTau217/Aβ42** | Normal (n=227) | 0.762 (0.693 - 0.831) | 78% (69% - 87%) | 79% | 73% | 38% | 94% |
| | | MCI (n=45) | 0.818 (0.679 - 0.957) | 71% (62% - 80%) | 68% | 88% | 37% | 96% |
| | **pTau217** | Normal (n=227) | 0.728 (0.655 - 0.801) | 78% (69% - 87%) | 78% | 75% | 33% | 96% |
| | | MCI (n=45) | 0.753 (0.597 - 0.909) | 67% (57% - 77%) | 65% | 75% | 32% | 92% |
| | **Aβ42/40** | Normal (n=227) | 0.779 (0.713 - 0.845) | 75% (66% - 84%) | 82% | 57% | 53% | 84% |
| | | MCI (n=45) | 0.838 (0.724 - 0.952) | 76% (67% - 85%) | 76% | 75% | 63% | 85% |
| Quanterix Simoa | **pTau217/Aβ42** | Normal | 0.77 (0.702 - 0.838) | 77% (68% - 86%) | 78% | 70% | 33% | 94% |
| | | MCI (n=45) | 0.779 (0.633 - 0.926) | 71% (62% - 80%) | 68% | 88% | 37% | 96% |

| | | | | | | | | |
|---|---|---|---|---|---|---|---|---|
| | **pTau217** | Normal (n=227) | 0.781 (0.714 - 0.849) | 79% (71% - 87%) | 80% | 72% | 41% | 94% |
| | | MCI (n=45) | 0.766 (0.617 - 0.916) | 67% (57% - 77%) | 64% | 83% | 26% | 96% |
| | **Aβ42/40** | Normal (n=227) | 0.659 (0.579 - 0.74) | 73% (64% - 82%) | 76% | 53% | 27% | 91% |
| | | MCI (n=45) | 0.84 (0.722 - 0.958) | 69% (59% - 79%) | 66% | 86% | 32% | 96% |

Abbreviations: AUC=Area Under the ROC Curve, CI=Confidence Interval, LOC=Loss Of Consciousness, MCI=Mild cognitive impairment, NPV=Negative Predictive Value, PPV=Positive Predictive Value, Sen=Sensitivity, spe=Specificity, TBI=Traumatic Brain Injury.
For each biomarker/biomarker ratio, performance metrics were derived using a threshold that optimizes its overall accuracy.

### Supplementary Table S5. Aβ40/42, pTau217and pTau217/Aβ42 performance vs SUVR threshold (1.17)

*Supplementary Table S5. Aβ40/42, pTau217and pTau217/Aβ42 performance vs. SUVR thresholds by TBI category*

| **Platform** | **Biomarker** | **TBI group** | **AUC (95% CI)** | **Accuracy (95% CI)** | **PPV** | **NPV** | **Sen** | **Spe** |
|---|---|---|---|---|---|---|---|---|
| Fujirebio Lumipulse | **pTau217/Aβ42** | None (n=93) | 0.834 (0.706 - 0.961) | 90% (84% - 96%) | 92% | 79% | 65% | 96% |
| | | TBI with LOC 0-5min (n=89) | 0.727 (0.573 - 0.882) | 84% (76% - 92%) | 89% | 60% | 53% | 92% |

*Supplementary Table S5. Aβ40/42, pTau217and pTau217/Aβ42 performance vs. SUVR thresholds by TBI category*

| Platform | Biomarker | TBI group | AUC (95% CI) | Accuracy (95% CI) | PPV | NPV | Sen | Spe |
|---|---|---|---|---|---|---|---|---|
| | | TBI with LOC >5min (n=90) | 0.747 (0.636 - 0.858) | 71% (62% - 80%) | 73% | 61% | 37% | 88% |
| | **pTau217** | None (n=93) | 0.807 (0.685 - 0.929) | 87% (80% - 94%) | 88% | 78% | 41% | 97% |
| | | TBI with LOC 0-5min (n=89) | 0.697 (0.533 - 0.86) | 86% (79% - 93%) | 87% | 78% | 41% | 97% |
| | | TBI with LOC >5min (n=90) | 0.708 (0.594 - 0.822) | 71% (62% - 80%) | 72% | 64% | 30% | 92% |
| | **Aβ42/40** | None (n=93) | 0.813 (0.715 - 0.91) | 80% (72% - 88%) | 84% | 43% | 18% | 95% |
| | | TBI with LOC 0-5min (n=89) | 0.789 (0.669 - 0.909) | 85% (78% - 92%) | 87% | 70% | 41% | 96% |
| | | TBI with LOC >5min (n=90) | 0.776 (0.679 - 0.873) | 75% (66% - 84%) | 75% | 79% | 37% | 95% |
| Quanterix Simoa | **pTau217/Aβ42** | None (n=93) | 0.834 (0.699 - 0.968) | 88% (81% - 95%) | 89% | 80% | 47% | 97% |

*Supplementary Table S5. Aβ40/42, pTau217and pTau217/Aβ42 performance vs. SUVR thresholds by TBI category*

| Platform | Biomarker | TBI group | AUC (95% CI) | Accuracy (95% CI) | PPV | NPV | Sen | Spe |
|---|---|---|---|---|---|---|---|---|
| | | TBI with LOC 0-5min (n=89) | 0.687 (0.547 - 0.827) | 82% (74% - 90%) | 86% | 55% | 35% | 93% |
| | | TBI with LOC >5min (n=90) | 0.791 (0.688 - 0.893) | 79% (71% - 87%) | 77% | 87% | 43% | 97% |
| | **pTau217** | None (n=93) | 0.847 (0.735 - 0.959) | 89% (83% - 95%) | 91% | 77% | 59% | 96% |
| | | TBI with LOC 0-5min (n=89) | 0.717 (0.562 - 0.871) | 84% (76% - 92%) | 88% | 62% | 47% | 93% |
| | | TBI with LOC >5min (n=90) | 0.782 (0.678 - 0.887) | 73% (64% - 82%) | 74% | 69% | 37% | 92% |
| | **Aβ42/40** | None (n=93) | 0.806 (0.71 - 0.902) | 82% (74% - 90%) | 87% | 50% | 41% | 91% |
| | | TBI with LOC 0-5min (n=89) | 0.629 (0.498 - 0.76) | 75% (66% - 84%) | 82% | 27% | 18% | 89% |
| | | TBI with LOC >5min (n=90) | 0.769 (0.652 - 0.887) | 73% (64% - 82%) | 73% | 71% | 33% | 93% |

*Supplementary Table S5. Aβ40/42, pTau217and pTau217/Aβ42 performance vs. SUVR thresholds by TBI category*

| Platform | Biomarker | TBI group | AUC (95% CI) | Accuracy (95% CI) | PPV | NPV | Sen | Spe |
|---|---|---|---|---|---|---|---|---|

Abbreviations: AUC=Area Under the ROC Curve, CI=Confidence Interval, LOC=Loss Of Consciousness, NPV=Negative Predictive Value, PPV=Positive Predictive Value, Sen=Sensitivity, spe=Specificity, TBI=Traumatic Brain Injury.
For each biomarker/biomarker ratio, performance metrics were derived using a threshold that optimizes its overall accuracy.

SUVR derived using methods described by Wiener et al., 2023

### Supplementary Table S6: Sensitivity analysis- Aβ42/40, pTau217 and pTau217/Aβ42 performance vs amyloid PET visual read by TBI category for participants whose TBI occurred at least 10 years ago

*S6: Aβ40/42, pTau217and pTau217/Aβ42 performance by TBI category in participants whose TBI occurred at least 10 years ago*

| Platform | Biomarker | TBI group | AUC (95% CI) | Accuracy (95% CI) | PPV | NPV | Sen | Spe |
|---|---|---|---|---|---|---|---|---|
| Fujirebio Lumipulse | **pTau217/Aβ42** | None (n=93) | 0.88 (0.8 - 0.97) | 89% (83% - 95%) | 90% | 82% | 53% | 97% |
| | | TBI with LOC 0-5min (n=75) | 0.71 (0.58 - 0.85) | 77% (68% - 86%) | 80% | 62% | 40% | 91% |
| | | TBI with LOC >5min (n=78) | 0.75 (0.65 - 0.86) | 64% (54% - 74%) | 62% | 71% | 34% | 88% |

*S6: Aβ40/42, pTau217and pTau217/Aβ42 performance by TBI category in participants whose TBI occurred at least 10 years ago*

| Platform | Biomarker | TBI group | AUC (95% CI) | Accuracy (95% CI) | PPV | NPV | Sen | Spe |
|---|---|---|---|---|---|---|---|---|
| | **pTau217** | None (n=93) | 0.86 (0.77 - 0.95) | 87% (80% - 94%) | 88% | 78% | 41% | 97% |
| | | TBI with LOC 0-5min (n=75) | 0.67 (0.53 - 0.82) | 77% (68% - 86%) | 78% | 71% | 25% | 96% |
| | | TBI with LOC >5min (n=78) | 0.72 (0.61 - 0.83) | 64% (54% - 74%) | 62% | 77% | 29% | 93% |
| | **Aβ42/40** | None (n=93) | 0.81 (0.68 - 0.93) | 79% (71% - 87%) | 91% | 46% | 65% | 83% |
| | | TBI with LOC 0-5min (n=75) | 0.81 (0.71 - 0.92) | 74% (65% - 83%) | 84% | 52% | 60% | 80% |
| | | TBI with LOC >5min (n=78) | 0.75 (0.64 - 0.86) | 71% (62% - 80%) | 68% | 77% | 49% | 88% |
| Quanterix Simoa | **pTau217/Aβ42** | None (n=93) | 0.88 (0.78 - 0.98) | 88% (81% - 95%) | 89% | 80% | 47% | 97% |
| | | TBI with LOC 0-5min (n=75) | 0.67 (0.54 - 0.81) | 74% (65% - 83%) | 78% | 55% | 30% | 91% |

*S6: Aβ40/42, pTau217and pTau217/Aβ42 performance by TBI category in participants whose TBI occurred at least 10 years ago*

| Platform | Biomarker | TBI group | AUC (95% CI) | Accuracy (95% CI) | PPV | NPV | Sen | Spe |
|---|---|---|---|---|---|---|---|---|
| | | TBI with LOC >5min (n=78) | 0.77 (0.66 - 0.88) | 65% (55% - 75%) | 62% | 79% | 31% | 93% |
| | **pTau217** | None (n=93) | 0.85 (0.73 - 0.97) | 87% (80% - 94%) | 89% | 73% | 47% | 96% |
| | | TBI with LOC 0-5min (n=75) | 0.75 (0.63 - 0.88) | 78% (69% - 87%) | 78% | 83% | 25% | 98% |
| | | TBI with LOC >5min (n=78) | 0.77 (0.66 - 0.87) | 63% (53% - 73%) | 61% | 75% | 26% | 93% |
| | **Aβ42/40** | None (n=93) | 0.81 (0.7 - 0.91) | 78% (69% - 87%) | 89% | 43% | 53% | 84% |
| | | TBI with LOC 0-5min (n=75) | 0.43 (0.29 - 0.58) | 66% (56% - 76%) | 76% | 37% | 35% | 78% |
| | | TBI with LOC >5min (n=78) | 0.7 (0.58 - 0.83) | 69% (59% - 79%) | 66% | 79% | 43% | 91% |

Abbreviations: AUC=Area Under the ROC Curve, CI=Confidence Interval, LOC=Loss Of Consciousness, NPV=Negative Predictive Value, PPV=Positive Predictive Value, Sen=Sensitivity, spe=Specificity, TBI=Traumatic Brain Injury.
For each biomarker/biomarker ratio, performance metrics were derived using a threshold that optimizes its overall accuracy.
